# Understanding sports betting as an emerging behavioral addiction among young men in Ethiopia

**DOI:** 10.64898/2026.10.01.26363633

**Authors:** Mesay Teshome, Beakal Amare, Awoke Mihretu

**Author notes:** Corresponding Author: Awoke Mihretu, P.O.Box: 9086, Addis Ababa, Ethiopia.

## Abstract

**Background:** Sports betting is one of the most popular, rapidly evolving, and attractive forms of gambling. It is associated with an increased risk of gambling-related harms like relationship difficulties, crime, and physical and mental health problems. Understanding the subjective experiences of sports betting is essential for accurately defining problematic betting and avoiding the over pathologization of the behavior. Hence, this study explored the perceptions and experiences of young men involved in sports betting in Addis Ababa, Ethiopia.

**Methods:** We employed a qualitative study design using a phenomenological approach. In-depth interviews were conducted with thirteen purposively selected participants in Addis Ababa. Participants were adults aged 18 years or older who had engaged in sports betting within the past 12 months. They were recruited at sports betting houses. All interviews were transcribed verbatim and translated into English. The data were then imported into OpenCode software and analysed using reflexive thematic analysis, in which themes were actively generated through an iterative and interpretative process by the research team.

**Findings:** The data resulted in five major themes: reasons for engagement in sports betting, unique and attractive features of sports betting, individuals experience of problematic betting, subjective harms due to sports betting. We found that young men engage in sports betting for reasons like financial interests, influence from peers and friends, and as a way of spending time and dealing with boredom. This study also identified several features that make sports betting more appealing, including ease of access, the possibility of gambling with a small amount of money, the benefit of avoiding confrontation, giving football fans the opportunity to gamble on something they like, enabling people to test their knowledge about the sport, the availability of betting options, and more rewards and reinforcements compared to other forms of gambling. Patterns suggestive of problematic betting include preoccupation and extensive planning, a tendency to bet successively and take more risky options after losing on a bet, unsuccessful attempts to quit, lying about involvement, social and occupational impairment, and negative emotional experiences.

**Conclusion:** Sports betting emerged in participants accounts as a growing behavioral practice among young men in Addis Ababa, Ethiopia, raising concerns for those who described their experiences While often initiated as a leisure activity, participants’ narratives suggested patterns that extended beyond casual engagement and shared features commonly associated with problematic gambling. Future studies could include women and use large-scale quantitative designs to explore the scope and distribution of sports betting-related harms at the population level and to help inform appropriate interventions.

## Background

Sports betting is the activity of predicting sports results and placing a wager on the outcome (Michael, 2020). According to the DSM-5 gambling involves risking something of value in the hopes of obtaining something of greater value and it is a more common activity among young and middle-aged people. DSM-5 also includes sports betting as a form of gambling preferred by younger group of individuals(APA, 2022).

Previous studies have identified some forms of gambling, such as sports betting are associated with increased risk for gambling related harms like relationship difficulties, crime, physical and mental health problems(McDaid, 2018). Although gambling disorder is a recognized diagnosis in the DSM-5, the specific characteristics that define problematic sports betting remain poorly documented.

The global prevalence of severe problematic gambling is estimated to range between 0.1% and 5.8% (Bijker R, 2022). This indicates a substantial need for support among individuals experiencing gambling-related difficulties. Research has further suggested that the harm associated with gambling can be comparable to that caused by major depressive disorder, substance use, and dependence (Fong, 2005).

Young adults engaged in problematic gambling behaviors are reported to be at an increased risk of developing major mental disorders (Afifi, 2016). Recent trends indicate that sports betting is increasingly becoming a part of everyday life in Africa, where opportunities for meaningful employment among young people are limited, making gambling appear as a viable means of earning income. In some African countries, such as Kenya, estimates suggest that up to 76% of the young population participates in gambling (Wangari, 2017).

Online sports betting is a relatively recent phenomenon in Ethiopia, yet it is already widespread and rapidly expanding (Abdi et al., 2015). European football commands a substantial fan base in the country, and strong team identification combined with perceived knowledge about matches may increase engagement in betting activities. As a result, the prevalence of both overall and problematic gambling is likely to rise in the foreseeable future. Given that the industry primarily targets young and economically productive segments of the population, this trend may have serious social and economic consequences at the national level(Afrika., 2026; Mihret & Eskezia, 2024).There are only a few cross-sectional surveys in Ethiopia that show the prevalence of sports betting but inductive exploration of the reasons for their engagement and how their practice is problematic is not systematically explored(Abdi et al., 2015). There are also controversies regarding the impact of sports betting as reflected in several media outlets in Ethiopia, several stakeholders presenting their opposing views on the matter(Sichali et al., 2023). There appears to be some confusion on the government’s side whether to license or close sporting betting business in the country. In addition, little is known about how sports betting is different from other forms of gambling. The perceived impact of sports betting on the lives of the bettors is also not well studied. Hence, this study aimed to explore the perceptions and experiences of young men involved in sports betting in Addis Ababa, Ethiopia.

## Materials and methods

### Study design

A phenomenological qualitative study design was employed as we explored the subjective experiences of individuals while they are engaged in online sports betting, their perception and how they might think sports betting can be harmful. Preference for online versus in-person sports betting appears to be driven primarily by levels of technological literacy rather than by betting patterns, such as the severity or problematic nature of gambling behavior.

### Study setting

The study was conducted around sports betting shops found in different parts of Addis Ababa, the capital city of Ethiopia, which represent their respective betting companies. There are more than ten online betting sites in Ethiopia licensed by the national lottery administration of Ethiopia. Different gambling brands designed their own cash deposit and withdrawal systems.

#### Sampling

A total of 13 participants were selected for the qualitative component based on the principle of theoretical saturation. We believe that this sample size was sufficient to generate a comprehensive understanding of the phenomenon and to adequately address the study aims.

We selected participants who were actively engaged in sports betting at the time of our visits, specifically those who were placing bets and in possession of betting tickets.

### Eligibility criteria

Individuals who were 18 or older, those who were able to participate in an interview conducted in Amharic language and those who were involved in sports betting for the past 12 months were eligible for the study.

### Data collection methods

A semi structured interview guide was developed, comprising open ended questions that covered key thematic areas relevant to the study objectives. The topic guide was prepared in Amharic and used consistently across interviews. On average, each interview lasted approximately 50 minutes. Interviews were conducted in private settings, either within participants’ home compounds or in quiet community locations to ensure confidentiality. All interviews were conducted by a psychiatry specialty trainee. Data management and analysis

After data collection, all interviews were transcribed verbatim following at least three careful listenings of the audio recordings. The transcripts were then translated from Amharic into English. The primary translator was a bilingual psychiatry trainee fluent in both Amharic and English. To ensure accuracy, the original Amharic transcripts and their English translations were reviewed and compared by co-authors. In addition, both versions were cross checked during the coding process to maintain conceptual equivalence. The finalized transcripts were imported into Open Code version 4.02, a software program designed for coding and categorizing qualitative data generated from interviews and observations. We followed the reflexive thematic analysis steps outlined by Virginia Braun and Victoria Clarke (2006) to analyze the data(Braun, 2006). Investigators conducted open coding by systematically coding relevant words, phrases, sentences, and analytic memos. Through iterative reading and constant comparison, codes were organized into subthemes and subsequently grouped into overarching themes. A comprehensive report of the thematic analysis was then produced. Any inconsistencies in translation or coding were resolved through discussion among the research team and, when necessary, through consultation with the senior co-author to reach consensus.

### Data quality assurance

After the participants were contacted in person, interviews were conducted in a safe, and quiet environment (to ensure the quality and accuracy of the study.) Senior co-authors also contributed with coding and identification of themes.

## Findings

### Socio-demographic characteristics of the Respondents

Participants were males, ranging in age from 22 to 37; all of them were single; seven of the participants were orthodox Christians. Most of the participants have been involved in betting for more than two years.

**Table 1:** Sociodemographic characteristics of participants.

| Code Number | Sex | Marital status | Religion | Number of years of involvement | Occupation |
| --- | --- | --- | --- | --- | --- |
| 001 | male | Single | Orthodox Christian | 3 years | Daily laborer |
| 002 | male | Single | Orthodox Christian | 1 year | Vocalist/Merchant |
| 003 | male | Single | Muslim | Six months | Security guard |
| 004 | male | Single | Muslim | 5 years | Shoemaker |
| 005 | male | Single | Orthodox Christian | 3 years | Unemployed |
| 006 | male | Single | Muslim | 5 years | Daily laborer |
| 007 | male | Single | Muslim | 3 years | Student |
| 008 | Male | Single | Orthodox<br>Christian | 3 years | Unemployed |
| 009 | Male | Single | Muslim | 3 years | Unemployed |
| 010 | Male | Single | Orthodox<br>Christian | 3 years | Carwash |
| 011 | Male | Single | Orthodox<br>Christian | 2 years | Driver |
| 012 | Male | Single | Orthodox<br>Christian | 1 year and half | Unemployed |
| 013 | Male | Single | Muslim | 2 years | Driver |

The findings are presented in the following four major themes: reasons for engagement in sports betting, unique and attractive features of sports betting, individual experiences of problematic betting and subjective harms due to sports betting.

### Reasons for engagement

#### Financial Interest

Many participants described sports betting as a means to improve their financial situation. The primary motivation was the hope of turning small amounts of money into large winnings:

> “I have no special feeling toward betting—just the idea of winning money. If I have 100 birr, I think about turning it into 300.” (Age 35-40, security guard)

Some equated betting with other desperate pursuits of economic change:

> “You’re young and want change—it’s like those who migrate abroad for a better life.” (Age 30-35, unemployed)

The affordability of betting makes it especially appealing in today’s economy:

> “I never thought of quitting. You could win 10,000 birr from a 10-birr bet. Ten birr is basically worthless these days.” (Age 30-35, daily laborer)

For some, this created dependency and diminished their motivation to work:

> “When you believe you might win big, working starts to feel pointless.”

#### Social Influence

Nearly all participants reported that their initiation into betting was influenced by peers. Friends introduced them to betting and modeled the behavior:

> “I started five years ago because of friends. They used to gamble on their phones and showed me how. Now I’m worse off than them. As they say, ‘You resemble your friends.’” (Age 30-35, shoemaker)

Betting was often framed socially as both entertainment and a potential income source:

> “They said it’s fun, a good way to pass time, and you can earn money.” (Age 20-25, vocalist)

#### Way of Spending Spare Time

Unemployment was frequently cited as a reason for betting—not just for the money, but as a way to fill time:

> “Most people you see betting are unemployed. If you had a job, you wouldn’t have time to stand around buying tickets.” (Age 35-40, daily laborer)

The availability of games on weekends and throughout the week made betting an accessible and engaging pastime.

### Unique and Attractive Features of Sports Betting

#### Convenience

Most participants highlighted the accessibility of sports betting—both in terms of low financial entry and ease of use:

> “Losing 10 birr means nothing now because of inflation. But you might win a million.” (Age 35-40, daily laborer)
>
> “You can place bets on your phone—no need to go to a shop.” (Age 30-35, shoemaker)

The digital nature of betting also reduced interpersonal conflicts:

> “Before, we bet face-to-face and fights could happen. Now you bet separately at a shop and avoid those issues.” (Age 30-35, unemployed)

#### Emotional Connection to Sports

Participants described betting on sports they already enjoyed—especially football—as a unique and emotionally engaging experience:

> “Betting on something you like, like football, makes you happy—and you might earn money too.” (Age 20-25, student)

Their knowledge of European leagues boosted confidence and made them more likely to bet:

> “We watch football, so we bet on football. Ask me to bet on basketball, and I wouldn’t— I’d be afraid of losing.” (Age 30-35, car washer)

Winning was not just financial—it was tied to a sense of competence:

> “Winning 200 birr from a bet feels better than getting 300 as a gift. It proves my prediction was right.” (Age 20-25, vocalist)

#### Low Effort

Betting was described as an easy way to make money, especially compared to traditional work:

> “It’s sweet money—you earn it without effort.” (Age 20-25, vocalist)
>
> “Unlike playing pool, which takes time and energy, a Keno game takes just three minutes.” (Age 30-35, security guard)

#### Variety and Multiple Options

Participants appreciated the wide range of betting options. Within a single match, there are many possible outcomes to bet on:

> “There are over 30 options for a single match—who scores, who gets a card, who’s sent off. You feel like, ‘Maybe this one will work.’” (Age 30-35, shoemaker)

They also mentioned betting features like “double chance,” which improves odds of winning:

> “Double chance means your team can win or draw—you’re covered either way.” (Age 25-30, unemployed)

#### Perceived Rewards

Most participants believed betting offers better returns than other forms of gambling:

> “You might win a lot of money from just 10 or 20 birr. That gets you excited.” (Age 25-30, unemployed)

Betting platforms also offer incentives like consolation prizes or refunds:

> “If I bet on four matches and get three right, I still get 80 birr. If I get two right, they refund my original bet. That makes me happy.” (Age 20-25, vocalist)

### Individuals experience of problematic betting

#### Being preoccupied with sports betting

Sports betting involves thinking a lot about finding ways of winning to the extent of preparing formulas. Those who are employed stressed the time-consuming nature of this experience and the negative impact it had on their work performance. Those who are currently unemployed reported it interfered with their ability to plan for the future and take job opportunities when they arrive.

> *“I’m a vocalist. The bets and numbers come to my mind while I’m singing in a club. The numbers pass in front of my eyes. Sometimes I see it in my dreams. There were occasions that I told my friends, “I saw the numbers, 50 birr, 1050 birr.”” (Age 20-25, vocalist)*

According to participants, while being a football fan can influence involvement in sports betting, placing bets on football matches further intensifies an individual’s interest in watching the games. *“I was fed-up thinking about football. It makes you think about football all the time. It makes you think about it all the time. While going and coming from work “(Age 25-30 unemployed)*

#### Chasing losses

Another habit reported by most of the participants is the tendency to place bets successively to win back the money lost on the previous bet. This is made possible by the availability of successive matches, especially on the weekends on which the person could go again.

> *“If I lose, I bet on the next game. You promise to yourself to improve what you missed on the previous game. The game its self creates an urge to bet frequently. You say things like I shouldn’t have predicted like this*.*” (Age 35-40, daily laborer)*

Participants mentioned the feeling one experiences after the loss as the main reason behind their subsequent bets and also forces them to entertain more risky options

*After losing you will become emotional and bet with bigger money in order to get back the money you lost. Because of your emotions and, you start to bet with bigger money and smaller matches. You increase the amount of money you bet and decrease the number of matches you have to wait to win the money. (Age 35-40, daily laborer)*

Another participant shared a similar experience of not letting the loss go

> *“After losing on a bet, you start to fight because you are a man. After losing all the money you brought, you start to ask you gambling friend for a loan. Since individuals who gamble tend help each other out by nature, he will give you some loan to try your luck. If you lose, you will leave the place (Age 30-35, unemployed)”*

Almost all participants mentioned this habit of “placing bets until you get it right’ is worse with the betting game called keno. With this game the results of the bets are displayed every two-three minute and the gambler has the opportunity to bet again within a very short interval.

> *“You may have to wait 90 min if you want to bet on a football game but with keno you can place bets almost every minute and it’s available almost 24 hours. It takes what you have got*.*” (Age 35-40, daily laborer)*

#### Unsuccessful attempts to quit

Participants described a desire to quit their involvement in sports betting and previous attempts to quit with variable degree of success. Some of them mentioned attempts to quit which didn’t last for more than few days. The temporary feeling of guilt and disinterest disappear as soon as one has the access to money.

> *“I thought I lost my desire. I used to watch movies at home with the time that I had. But after getting money I start to play again as a fresh gambler. Isn’t this an addiction on its own?” (Age 35-40, security guard)*

The same participant shared his difficulty to quit by comparing it to other forms of addiction.

> *“I’m dependent on it. I don’t have a good attitude towards it. Its impact on people’s lives is like tobacco addiction. Just like you smoke knowing its impacts on your health, you gamble though you know it is harming you*.*” (Age 30-35, shoemaker)*

Another participant shared the experience of difficulty to quit because of his continued involvement in watching football. The game is a reminder for betting.

> *“I tried multiple times. But the things that I told you, there is an influence of watching the game. For example, I remember betting when I watch games. That made it difficult for me to quit*.*” (Age 30-35, car wash)*

#### Lie about involvement in betting

Most of the participants reported they lie about their involvement in sports betting in one way or another. The lie is towards family members most of the time or friends who are critical of the behavior.

*It had an impact on my social life. First it makes me lie. I lie because it is a shameful place to be and I tell people “I’m somewhere else” when they call me. (Age 30-35, unemployed)*

### Subjective harms due to sports betting

#### Financial Harm

Participants consistently emphasized that most of the harm caused by betting stemmed from financial loss. They described how these losses triggered emotional distress, strained family and social relationships, and contributed to a general sense of stagnation.

> “For me it is very harmful. For example, now I brought the money that I was saving in my account. I do this thing usually. It is like, the fact that I’m working has no value if I waste my money on this.” (Age 20-25, vocalist)

Financial loss impacted everything from leisure activities to essential needs.

> “If you have a job, Sunday is your off day. If you lose your money on a bet, you don’t have anything to go out and chill with.” (Age 30-35, car wash)
>
> “We sold a printer. What did we do with the money? We are betting with it. You may come here and play with the money you were supposed to buy shoes with.” (Age 20-25, vocalist)

#### Negative Emotional Experiences

Sports betting was associated with a wide range of negative emotions that affected mental wellbeing. Guilt was frequently mentioned—often due to money lost, time wasted, or religious conflicts.

> “I spend more than five hours per day. Maybe it’s something that foreign industries brought to steal our time.” (Age 35-40, security guard)
>
> “I’m a Muslim… I’m involved in betting knowing that it’s forbidden.” (Age 30-35, shoemaker)
>
> “From a religious perspective… this is like pushing His will aside. I feel bad when I pray.” (Age 20-25, unemployed)

Anger was another recurring emotion, especially after near-wins.

> “Once I placed a bet on Manchester City. They won 14 matches and lost the final. I was going to win 17,000 birr. I couldn’t fall asleep.” (Age 35-40, daily laborer)
>
> “Losing on a bet makes me angry. I may not reply to greetings properly.” (Age 20-25,vocalist)
>
> “It makes you very angry. It won’t go out of your mind for days.” (Age 20-25, unemployed)

Depression was reported after losses, especially when participants reflected on unmet financial needs.

> “I feel depressed on the day of the loss. You lose interest in talking, going out… You prefer to sleep.” (Age 30-35, car wash)
>
> “You won’t get along with others. You spend your time alone to avoid conflict.” (Age 25-30, unemployed)

Some participants experienced feelings of inferiority, linked to poor financial management.

> “I don’t have dependents. I should have saved more. People earning less live better than me.” (Age 30-35, shoemaker)

These emotions were often accompanied by a sense of exploitation by betting companies. Participants believed the system was designed to keep them losing.

> “Even those who claim to have won a lot have lost many times. The amount you lose is more than what you win.” (Age 35-40, daily laborer)

A few participants saw winning not just as financial gain but as revenge against the system.

> “When I win, I feel happy. It’s like revenge. They’re taking my money—when I get them back, it feels good.” (Age 20-25, vocalist)

#### Social and occupational impairment

Participants also mentioned the negative impacts their involvement in sports betting had on their motivation and performances at work. Difficulty getting along with people at work including customers was one of the incriminated reasons presented by the participants

> “*After losing, on the following day, I feel down and angry. It interferes with your work. I wash cars and I may not treat my customers properly” (Age 30-40, car wash)*

Wasting time was another factor mentioned to interfere with their jobs

*It has some impact. I’m a vocalist, and I may not practice as long as I should. It takes some of my time. (Age 20-25, vocalist)*

Participants also mentioned early departure from work to buy the tickets and follow matches

> *“For example, if there is a match today, it’s a must to stop working and buy the tickets. That part interferes with your work” (Age 30-35, car wash)*

The prospect of winning money like their friends was mentioned as the reason behind their lack of motivation.

> *“The feeling I have right now, the Motivation I had for work had declined significantly from previous times. I think most individuals feel like this. The hope of getting money this way decreases your motivation for work*.*”(Age 35-40, security guard)*

Interpersonal relationship is another area affected by wasting time due to betting (as it takes, they time they were supposed to give for their families and friends)

> *“You may not have the time for your friends. There may also be a conflict. I remember occasions where I forgot I had to meet my friend; he usually calls and yells at me” (age 20-25, vocalist)*

Similar to occupational performance their relations with others is also affected by the negative emotions they experience after losing on a bet or things going against them

> *“There are times where I would argue with my family members while waiting for the results of the bets I placed. If things are not going my way, I tend to argue and fight with my family which is not my normal way of behaving*.*” (Age 30-35, shoemaker)*

Participants also reported their families do not like their involvement in betting and there is a conflict because of it

> *“Your family does not want stuff like this. Since they opened the shops everywhere you are likely to be seen by one of your family members. They say to me why are you involved in stuff like this? I reply don’t bother me; I’m playing with my own money. They are not happy about it. (Age 30-35, shoemaker)”*
>
> *“Sometimes I come home very late at night, like I said, your mood might not be right after you lose and you start to talk to yourself starring at the moon. You don’t want your family to see your change in mood. You stay outside until that goes away*.*” (Age 25-30, unemployed)*

An individual’s affiliation to betting changes their attitude towards money and makes them greedy according to the participants. Friendship is all about reaching out for one another at times of need and crisis. The affection one develops for money forces the gambler to lose this connection.

> *“When you friend is in need he asks for a loan. Betting takes that away. It makes you greedy. You want from others instead of giving. You will have no real friendships. The game becomes your friend*.*” (Age 30-35, unemployed)*

## Discussion

This study aimed at exploring the reasons individuals engage in those behaviors, what kinds of harm they sustain, and what problematic patterns they constitute. The study identified four major themes: reasons for involvement in sports betting, attractive features of sports betting, subjective harms due to sports betting, and individual’s experiences of problematic betting.

Participants in this study mentioned several reasons for their engagement in sports betting, like financial interests, including a means of earning money and changing one’s life, hearing attractive and dramatic stories about people winning huge amounts of money, influence from their peers and friends, and a way of spending time to mitigate the boredom associated with unemployment. Another qualitative study conducted on 17 young individuals reported the importance of various factors in people’s involvement in in-play betting, like its association with more excitement, making the game more intense, and allowing the gambler to use his gambling skills(Elizabeth A Killick & Mark D Griffiths, 2021). They also emphasized the importance of the cash-out features because they allow the individual to recoup from losing a bet.

In this particular study according to the majority of the respondents unique and attractive features of sports betting include its convenience (including the ease of access one has via his/her mobile phone, allowing a person to gamble with a small amount, and the fact that it allows gamblers to avoid direct confrontation), its interesting nature in the sense that it allows individuals to place a bet on something they like (football for most of the participants), and enables them to test their knowledge about the sport, the availability of multiple ways to bet which increases the perceived chance of winning and its potential rewards (the prospect of winning a lot of money for a very small wager). Which is consistent with other studies which identified the ease of access one has, the fact that it is becoming normalized, and having many attractive features that may contribute to involvement in sports betting(Etuk et al., 2022).

Consequences that the participants perceive as harmful outcomes of their involvement in sports betting include psychosocial harms like experiencing guilt resulting from the financial implications and contradicting their religion, anger, and feeling down, which affect their interpersonal relations and occupational performance. According to the DSM-5, jeopardizing relationships or career opportunities due to gambling is among the least often endorsed criteria and most often occurs among those with more severe gambling disorders(Association, 2013). Loss of self-respect and a sense of inferiority because of their failure to meet certain expectations were also among the findings of this study. In other studies, problematic online sports betting, as compared to non-problematic online sports betting, was associated with more psychological and emotional distress (Valenciano-Mendoza et al., 2023). Regarding an individual’s experience of problematic betting, most of the participants reported preoccupation, which mostly takes the form of extensive planning regarding possible ways of winning, chasing losses as shown by the tendency to bet successively and taking more risky options after a losing bet, a desire to stop in most cases but unable to do so due to peer and environmental influences, and lying about their involvement in sports betting to families and friends due to the shame associated with participation in gambling.

The themes emerging from the current study are also closely align with findings from similar research conducted in the Global North(E. A. Killick & M. D. Griffiths, 2021). Many of the features that make sports betting particularly attractive, compared with other forms of gambling, are common across different contexts. For instance, the use of the “cash-out” feature, which generates heightened excitement, and the bettors’ illusion of control—referred to in this study as *mind preoccupations*—mirror the “online sports betting loop” described by Parke and Parke (Parke & Parke, 2019).

According to the DSM-5, the most frequently endorsed criteria for gambling disorder are related to preoccupation with gambling and “chasing” losses, which were also found among most of the participants in this particular study(Association, 2013). Similar to findings in this research, DSM-5 points out that a pattern of “chasing one’s losses” may develop, with an urgent need to keep gambling (often with the placing of larger bets or the taking of greater risks) to undo a loss or series of losses. The other findings of this study, including the desire to stop without succeeding and lying to conceal the extent of involvement, were also among the problematic behaviors diagnostic of a gambling disorder.

### Limitations

Despite concerted efforts by the study team, we were unable to recruit female participants. This challenge reflects broader contextual issues in Ethiopia, where high levels of stigma have similarly limited the inclusion of women in prior studies on behavioral addictions and substance use. Comparable recruitment challenges were also encountered for married individuals and participants from higher socioeconomic backgrounds. To enhance diversity, participants were purposively recruited from sports betting shops located in different areas of Addis Ababa. In addition, the inherent limitations of qualitative research including limited generalizability and reliance on self-reported accounts should be considered when interpreting the findings. Conclusions

Sports betting emerged in participants’ accounts as a growing behavioral practice among young men in Addis Ababa, Ethiopia, raising concerns for those who described their experiences. While often initiated as a leisure activity, participants’ narratives suggested patterns that extended beyond casual engagement and shared features commonly associated with problematic gambling. These included preoccupation with betting and deliberate planning, repeated or consecutive betting, substantial time investment, continued involvement despite reported social, financial, and psychological difficulties, increased risk-taking following losses, unsuccessful efforts to reduce or stop betting, and concealment of betting activities from family and friends.

The findings further highlight the need for further research to better understand sports betting within this context. Future studies could include women and employ large-scale quantitative designs to examine the scope and distribution of sports betting-related harms at the population level. Building on the present qualitative findings, in-depth and mixed-methods research may provide additional insight into pathways, meanings, and consequences of sports betting. Participants’ accounts also suggest that early, contextually appropriate interventions may be worth exploring.

## Data Availability

All data produced in the present study are available upon reasonable request to the authors

## Acknowledgements

The authors acknowledge the contributions of study participants. We would also like to acknowledge Addis Ababa University. Awoke Mihretu was supported through AMARI-II, funded through the DELTAS Africa Initiative (DEL-22-004).

## Author contributions

M.T. designed the study and A.M. and B. A supervised the study. M.T. contributed to data collection and analysis. M.T. and A.M. drafted the manuscript with supervision from B.A. All authors made critical intellectual contributions, read and approved the final manuscript and agreed to submission for publication.

## Data availability

The data that support the findings of this study are available from the corresponding author, A.M., on reasonable request.

## Declarations Ethical Approval

This study received ethical approval from the Research and Ethics Committee of the Department of Psychiatry, Addis Ababa University (MF/PSY/24/12).

## Consent to participate

Written informed consent was obtained from all participants after the objectives and procedures of the study were clearly explained. The study was conducted in strict accordance with the principles of the Declaration of Helsinki. No participant names were collected; however, addresses and other relevant information were obtained with explicit permission to facilitate follow up, and confidentiality was maintained throughout the study. All data were securely stored, and analyses were conducted anonymously.

## Consent for publication

Not applicable.

## Clinical trial number

Not applicable.

## Reference

Abdi, T. A., Ruiter, R. A., & Adal, T. A. (2015). Personal, social and environmental risk factors of problematic gambling among high school adolescents in Addis Ababa, Ethiopia. Journal of Gambling Studies, 31, 59–72.

Afifi, N. (2016). A longitudinal study of the temporal relationship between problemgambling and mental disorders amnog young adults. the canadian journal of psychiatry.

Afrika., i. (2026). Ethiopian iGaming market reshaped after shutdown in mid-December. iGaming Afrika. Retrieved February 2026, from https://igamingafrika.com/ethiopian-igaming-market-reshaped-after-shutdown-in-mid-december/. This article describes how the Ethiopian Lottery Service revoked all sports betting licences, causing a major market contraction, showing how widespread the sector had become before regulatory action.

APA. (2022). Diagnostic and statstical manual of mental disorders.

Association, A. P. (2013). Diagnostic and statistical manual of mental disorders (5 ed.) American Psychiatric Association.

Bijker R B. (2022). Global prevalence of help-seeking for problem gambling: A systematic review and meta-analysis. society for the study of addiction

Braun, V. (2006). Using thematic analysis in psychology. qualitative research in psychology. 77–101.

Etuk, R., Xu, T., Abarbanel, B., Potenza, M. N., & Kraus, S. W. (2022). Sports betting around the world: A systematic review. Journal of Behavioral Addictions, 11(3), 689–715.

Fong, T. W. (2005). The biopsychosocial consequences of pathological Grambling

Killick, E. A., & Griffiths, M. D. (2021). Why do individuals engage in in-play sports betting? A qualitative interview study. Journal of Gambling Studies, 37(1), 221–240.

Killick, E. A., & Griffiths, M. D. (2021). Why Do Individuals Engage in In-Play Sports Betting? A Qualitative Interview Study. J Gambl Stud, 37(1), 221–240. 10.1007/s10899-020-09968-9

McDaid, D. (2018). messuring gambling related-harms ; a framework for action. LSE Reaserch online.

Michael. (2020). sports betting. https://en.wikipedia.org/wiki/Sports_betting

Mihret, A. L., & Eskezia, A. T. (2024). Psychosocial and economic impacts of betting among the youth and adults in Bahir Dar City, Ethiopia: Impact of betting. Ethiopian Journal of Social Sciences, 10(2), 104–124.

Parke, A., & Parke, J. (2019). Transformation of sports betting into a rapid and continuous gambling activity: A grounded theoretical investigation of problem sports betting in online settings. International Journal of Mental Health and Addiction, 17(6), 1340–1359.

Sichali, J. M., Bunn, C., McGee, D., Marionneau, V. K., Yendork, J. S., Glozah, F., Udedi, M., & Reith, G. (2023). Regulation of gambling in Sub-Saharan Africa: findings from a comparative policy analysis. Public Health, 214, 140–145.

Valenciano-Mendoza, E., Mora-Maltas, B., Mestre-Bach, G., Munguía, L., Richard, J., Derevensky, J. L., Potenza, M. N., & Jiménez-Murcia, S. (2023). Clinical correlates of sports betting: A systematic review. Journal of Gambling Studies, 39(2), 579–624.

Wangari, N. (2017). https://www.geopoll.com/blog/mobile-gambling-among-youth-in-sub-saharan-africa

